# Assessing and quantifying gait deviations in STXBP1-related disorder using three-dimensional gait analysis

**DOI:** 10.64898/2026.03.02.26346982

**Authors:** Merel Swinnen, Lis Gys, Kim Thalwitzer, Axel Deporte, Charlotte Van Gorp, Eileen Vermeer, Firooz Salami, Sarah Weckhuysen, Sebastian I. Wolf, Steffen Syrbe, An-Sofie Schoonjans, Ann Hallemans, Hannah Stamberger

**Author notes:** **Corresponding author:** Hannah Stamberger. shared first author (Merel Swinnen, Lis Gys).

## Abstract

**Background and objectives:** STXBP1-related disorder (STXBP1-RD), caused by pathogenic variants in the *STXBP1* gene, is a rare neurodevelopmental condition, characterized by early-onset seizures, developmental delay, intellectual disability (ID), and prominent motor dysfunction. Despite the high prevalence of motor symptoms, systematic gait characterization remains limited. We therefore aimed to quantitively assess gait in individuals with STXBP1-RD.

**Methods:** In this cross-sectional study, we included ambulatory patients aged 6 years or older with genetically confirmed STXBP1-RD. Instrumented 3D Gait Analysis (i3DGA) was performed to objectively quantify gait. Functional mobility was assessed with the Functional mobility scale (FMS) and Mobility Questionnaire 28 (MobQues28). Caregiver health-related quality of life was evaluated using the PedsQL-Family Impact Module (PedsQL-FIM). We explored associations between gait, functional mobility, *STXBP1*-variant type and clinical features (ID, age at seizure onset, seizure frequency, age at onset of independent walking). Correspondence between i3DGA and the Edinburgh Visual Gait Score (EVGS), an observational gait assessment, was investigated.

**Results:** Eighteen participants were included. Compared to typically developing peers, individuals with STXBP1-RD had significantly reduced walking speed, step and stride length. Gait patterns were highly variable, with the most frequent pattern being an externally rotated foot progression angle (FPA), present in 11/18 participants. At home, 93.75% of the participants (16/18) walked independently, yet community mobility was more variable: 11/16 (68.75%) walked independently, 2/16 (12.50%) with aid and 3/16 (18.75%) used a wheelchair, indicating increasing limitations with distance and environmental complexity. Earlier acquisition of independent walking strongly predicted later unassisted ambulation at community level (p<0.001). Median MobQues28 score was 57.14% and median PedsQL-FIM score was 60.42%, indicating a moderate level of mobility limitations and reduced health-related quality of life of caregivers. EVGS was highly positive correlated with i3DGA (p= 0.001).

**Discussion:** Quantitative gait analysis in individuals with STXBP1-RD demonstrates heterogenous kinematic deviations, with an externally rotated FPA emerging as the most common pattern. Age at independent walking was a clinically relevant predictor of later functional mobility. EVGS showed strong correspondence with i3DGA and may offer a more practical, semi-quantitative assessment for broader use. These findings inform clinical decision-making and guide the selection of scalable outcome measures for natural history studies and interventional trials.

## 1. INTRODUCTION

Syntaxin-Binding Protein 1 (STXBP1), also known as Munc18-1, is a protein encoded by the *STXBP1*-gene that plays a key role in synaptic transmission in the brain (1, 2). Pathogenic variants in the *STXBP1* gene are associated with a neurodevelopmental phenotype now commonly referred to as STXBP1-related disorder (STXBP1-RD). Core clinical features of the disorder include (therapy resistant) epileptic seizures with mostly early-life onset, accompanied by developmental delay, intellectual disability and behavioural, communication and motor problems (3, 4). Importantly, there is significant phenotypic variability, with considerable differences in symptom profiles and severity between individuals (3–5). Although STXBP1-RD is considered a developmental and/or epileptic encephalopathy (DEE), the contribution of epileptic activity to developmental outcome remains difficult to delineate (6, 7).

In the motor development domain, gross motor milestones are typically delayed, and independent walking, a key milestone for motor independency, is only acquired in 30-50% of patients (6, 7). Motor problems frequently observed in STXBP1-RD include hypotonia or hypertonia (spasticity), and different types of movement disorders such as tremor, dystonia and ataxia. Furthermore, musculoskeletal abnormalities such as scoliosis and abnormal positioning of the feet can be present (3–5, 7). These issues may directly impair walking ability and contribute to the emergence of gait deviations in STXBP1-RD, although this has not been systematically quantified. Importantly, 30-50% of individuals with STXBP1-RD remain non-ambulatory, reflecting the extensive phenotypic heterogeneity of the disorder (5, 6). Interestingly, several studies have shown that earlier age at onset of seizures correlates with poorer motor outcomes, including reduced likelihood of achieving the ability to walk independently (5–7), whereas individuals without epilepsy tend to have milder motor impairment (7).

Functional mobility encompasses how individuals navigate their environment to perform daily activities, engage socially, and participate in community life (8, 9). It therefore reflects not only motor functioning but also constitutes a key determinant of social participation and interaction, with potential implications for the health-related quality of life of both patients and their caregivers (10, 11). Despite the high prevalence and presumed impact of gait and mobility problems in STXBP1-RD, systematic and quantitative gait characterization remains scarce.

Instrumented 3D Gait Analysis (i3DGA) enables detailed biomechanical characterization of gait, and has been used to identify characteristic gait patterns in other DEEs such as Dravet Syndrome (DS), where i3DGA distinguished two prevalent gait-patterns (12, 13). In DS, the severity of gait impairment correlates with functional mobility, underscoring the clinical relevance of quantitative gait assessment (14). Gait indices, such as the Gait Profile Score (GPS), derived from the i3DGA, offer a sensitive tool to objectively quantify gait-pathology across multiple joints and movement planes (14–18). However, its application is limited by the need for specialized gait analysis equipment, trained physiotherapists, and participants’ physical attendance at a fully equipped gait laboratory. Video-based gait assessment tools, such as the Edinburgh Visual Gait Score (EVGS), offer a more accessible, semi-quantitative alternative, particularly valuable for individuals with neurodevelopmental disorders such as STXBP1-RD, where applying i3DGA can be challenging (19).

## 2. MATERIALS AND METHODS

### 2.1 Study design

This cross-sectional study was part of a larger project investigating gait characteristics in rare genetic diseases (https://clinicaltrials.gov/study/NCT05161494). Results are reported according to the STROBE (Strengthening and Reporting of Observational Studies in Epidemiology) guidelines (20).

### 2.2 Participants

Participants were enrolled through the Antwerp University Hospital (Belgium) and the Heidelberg University Hospital (Germany). Eligible participants had a genetically confirmed diagnosis of STXBP1-RD and were aged 6 years or older. Participants were excluded if they were unable to walk unsupported or had a convulsive epileptic insult within 24 hours prior to the examination.

### 2.3 Standard Protocol Approvals, Registrations, and Patient Consents

This study was approved by the Ethics Committee of Antwerp University Hospital (Edge 002290, BUN B3002021000284). Approval for inclusion of clinical data from the German cohort was approved by Heidelberg University (S-320/2024). The privacy and security of personal patient information were ensured. Written informed consent for participation and publication was obtained for all participants by their legal guardians.

### 2.4 Data collection and analysis

Data collection took place from May 2023 until March 2025. Data was collected in two separate study visits, one at the neurology department to assess clinical (neurological) characteristics and one at the gait laboratory to assess gait, functional mobility and QoL.

### 2.5 Genetic and clinical characteristics

Following clinical characteristics of participants were retrieved from the medical records or obtained during the study visit with the (paediatric) neurologist: motor milestones including age of independent walking (in months), degree of ID according to the DSM-5 criteria (21), age at seizure onset (in months), seizure history (yes/no), seizure frequency (seizure free/yearly/weekly/daily) and the *STXBP1*-variant type. Current gross motor abilities were classified using the GMFCS (22).

### 2.6 Gait

i3DGA was performed at the M²OCEAN Movement Analysis Laboratory (University Hospital Antwerp) and the Heidel-MotionLab (Heidelberg University Hospital). To ensure comparability between sites, both laboratories used the same equipment and marker set; data processing steps were harmonized across sites. Participants performed a standard clinical gait analysis, barefoot at a self-selected pace until at least four representative valid left and right strides were collected. The Plug-in-Gait marker set-up was used and marker-movement was captured using a VICON system (10 to 12 camera’s, T10 or Vero 2.2, 100 Hz., Oxford Metrics, Oxford, UK). Segmental (pelvis and foot) and joint (hip, knee and ankle) kinematic profiles were calculated using Vicon Nexus Software (version 2.10.3). Based on the kinematic profiles of the separate trials, the GPS was calculated for each participant using MATLAB based on established methodology (15). For Heidelberg participants, GPS was calculated on the averaged walking trials. GPS was derived from 9 key kinematic variables: pelvic and hip kinematics in the sagittal, frontal, and transversal planes; knee and ankle kinematics in the sagittal plane; and the foot progression angle (FPA) in the transversal plane. GPS was expressed in degrees, with higher values indicating greater deviation from a normal gait pattern. The Movement Analysis Profile (MAP) decomposes the GPS into its 9 kinematic variables and provides a more detailed overview of the specific gait-deviations (15). Outcome of interest were GPS and MAP.

### 2.7 Functional mobility

Functional mobility was assessed using the parent-reported Functional Mobility Scale (FMS) and Mobility Questionnaire 28 (MobQues28). The FMS evaluates how the child moves in daily life at home (5 m; FMS-5), school (50 m; FMS-50), and in the community (500 m; FMS-500), scoring each distance from 1 (wheelchair) to 6 (independent walking on all surfaces) (9). The MobQues28 assesses performance of 28 daily motor activities on a 5-point scale from 0 (impossible without help) to 4 (not difficult at all). A total score was calculated as the sum of item scores divided by the maximum possible score, multiplied by 100, with higher values indicating better functional mobility (23). Outcome of interest were FMS-5, FMS-50, FMS-500 and MobQues28-total score.

### 2.8 Quality of Life

Health-related quality of life of one primary caregiver was assessed using the 36-item PedsQL-Family Impact Module (PedsQL-FIM), which evaluates eight domains: physical, emotional, social and cognitive functioning, communication, worry, daily activities, and family relationships. Items are rated on a 5-point scale (0 = never a problem, 4 = nearly always a problem), reverse-scored, and linearly transformed to a 0-100 scale, with higher scores indicating better caregiver quality of life and family functioning (24). Outcome of interest was the PedsQL-FIM-total score.

### 2.9 EVGS

Video recordings in frontal and sagittal planes were used for observational gait assessment and scored according to the 17 items of the EVGS. All analyses were performed by the same physiotherapist for all participants, blinded to GPS results at the time of assessment. Items were rated 0-2 (normal (0), moderate deviation (1), severe deviation (2)). In case of inconsistent scores across steps/videos, the mean of 2 scores was used. A total score (0–68) was calculated by summing left and right-side item scores, with lower score indicating less gait-pathology (eTable 2 in the supplemental files for scoring details). Due to GDPR-restrictions, videorecordings from Heidelberg could not be shared for EVGS scoring. Outcome of interest was the EVGS total score.

### 2.10 Statistical Analysis

Statistical analyses were performed using SPSS statistics software (IBM SPSS Statistics 31). Reported tests were performed 2-tailed with an alpha level for significance of p < 0.05, after adjustment for testing multiple endpoints, if applicable. Participants with missing values for variables of interest were excluded from the respective analyses. No imputation procedures were applied.

#### 2.10.1 Genetic and clinical characteristics

Descriptive statistics include median, minimum and maximum value for continuous variables (age at seizure onset, age of independent walking) and frequency counts for categorical variables (*STXBP1* variant type, seizure frequency, ID, GMFCS).

#### 2.10.2 Gait

Spatiotemporal parameters were compared to an age-matched control group of 18 individuals with typical gait, using a Mann-Whitney U test.

GPS and MAP were evaluated against a reference dataset comprising 60 typically developing individuals, aged between 4.6 and 18.6 years. We used the same methodology as reported in a previous study (14). Based on this comparison, individuals were classified into three categories: *normal* (≤ mean + 1SD), *mildly impaired* (> mean + 1SD) *and severely impaired* (≥ mean + 2SD). Median and interquartile range (IQR) are reported.

#### 2.10.3 Functional mobility and Quality of Life

Descriptive statistics include median and IQR for continuous variables (MobQues28 and PedsQL-FIM) and frequency counts for categorical variables (FMS).

#### 2.10.4 Correlations between GPS, functional mobility, genetic and clinical features

Spearman rank correlation was used for GPS and FMS-500. FMS-500 was selected for its greater inter-individual variability, relevance to mobility in community settings and easier interpretation than the Mobques28. FMS-500 was treated as an ordinal variable. Pearson correlation coefficient was applied for GPS and MobQues28 after confirming normality.

Correlation between GPS or FMS-500 and predefined genetic and clinical characteristics were subsequently explored. Mann-Whitney U tests assessed differences according to ID severity and *STXBP1*-variant type. Spearman rank correlations evaluated relationships with age at independent walking, age at seizure onset (restricted to individuals with seizure history) and seizure frequency. Seizure frequency was treated as an ordinal variable (0 = never experienced seizure, 1 = seizure free, 2 = yearly, 3 = weekly, 4 = daily). Bonferroni correction was applied for each statistical test type separately (α’ = 0.0125 for Mann-Whitney U tests; α’ = 0.0083 for Spearman correlations). Correlation strength was interpreted as negligible (0.00-0.30), low (0.30-0.50), moderate (0.50-0.70), high (0.70-0.90) or very high (0.90-1.00) (25).

#### 2.10.5 Correlation i3DGA and EVGS

The Pearson correlation coefficient was calculated between total GPS and EVGS total score to compare both evaluations, after testing for normality.

### 2.11 Data Availability

De-identified data not provided in the article can be made available by request from a qualified researcher.

## 3. RESULTS

### 3.1 Study population - Clinical characteristics

In total, 18 participants (age range 6-39 years), 11 females and 7 males, with a confirmed diagnosis of STXBP1-RD were enrolled in the study. Seven individuals carried an *STXBP1* missense variant, 7 a protein truncating variant and 4 a splice-site variant. Five participants never experienced seizures while 13 had a history of epilepsy. The median age at seizure onset was 4.5 months (range 1 - 96 months), and in 8 participants seizures started within the first year of life. Five participants (5/13) were seizure-free at the time of inclusion (no reported seizures within the past 2 years) and 8/13 still had active epilepsy with reported seizure frequencies ranging from daily seizures in 1 patient, weekly seizures in 4 and yearly seizures in 3.

All participants had some level of ID, 14 were categorized as severe to profound, 4 as mild to moderate ID. Information on motor development was available for 17/18 participants. Thirteen acquired independent walking after the age of 17.6 months, the 99^th^ percentile for healthy children according to WHO (26), with median acquisition age at 48 months (range 12 - 144 months). The GMFCS was determined in all participants, 5 individuals had a score of 1, 8 a score of 2 and 5 had a score of 3. Clinical and genetic characteristics per participant are reported in eTable 1 (in the supplemental files).

### 3.2 Gait

#### 3.2.1 Spatiotemporal parameters

Spatiotemporal parameters are reported in Table 1 and were compared to 18 age-matched typically developing peers (TD). STXBP1 participants had a decreased walking speed, step length and stride length compared to TD. Mean step width did not differ significantly, yet the STXBP1 cohort demonstrated greater variability than the TD group.

**Table 1:** Spatiotemporal parameters. STXBP1 vs. TD using Mann-Whitney U test, with significant result in bold.

| Spatiotemporal parameters | STXBP1 | TD | STXBP1 vs. TD |
| --- | --- | --- | --- |
|  | Median [Q1 - Q3] | Median [Q1 - Q3] | *p* |
| Gait speed (m/s) | 0.84 [0.63 - 1.07] | 1.24 [1.19 - 1.33] | **<0.001** |
| Step Length (m) | 0.46 [0.38 - 0.51] | 0.64 [0.56 - 0.68] | **<0.001** |
| Stride Length (m) | 0.93 [0.75 - 1.02] | 1.29 [1.13 - 1.36] | **<0.001** |
| Step Width (m) | 0.16 [0.12 - 0.28] | 0.14 [0.12 - 0.16] | 0.162 |
| Cadence (steps/min) | 109 [95 - 123] | 116 [111 - 131] | 0.109 |

#### 3.2.2 GPS and kinematic gait-profiles

The median score of the total GPS in our cohort was 10.2° (IQR 7.3° - 12.9°). Four participants (PT 4, 11, 12, 17) showed a normal total GPS, one (PT 7) gait was mildly impaired, and the remaining 13 participants showed severely impaired gait. At the group level, variation in GPS in our STXBP1 cohort did not reach statistical significance compared to typically developing peers. However, GPS in the study cohort showed broad variability. Across the movement planes, the most prominent variation was seen in the **transverse plane** (median 10.5°, IQR 7.2° - 15.7°). This was mostly driven by the *foot progression angle*, with deviations from typical gait ranging from 5.0° to 35.4°. Foot progression was more externally rotated in 11 participants either bilaterally (PT 3, 8, 9, 10, 14, 16, 18) or unilaterally (PT 1, 4, 6, 15). The smallest GPS variability was observed in the **frontal plane** (median 4.0°, IQR 3.1° - 4.8°) except for 1 extreme outlier (PT 10) who exhibited severely increased hip abduction and broad-based gait (step width 42.8 cm). GPS showed moderate variability in the **sagittal plane** (median 9.0°, IQR 6.6° - 12.8°). Nine participants showed increased *hip flexion/extension* scores, bilaterally (PT 1, 2, 10, 13, 15, 18) or unilaterally (PT 7, 16), attributable to a reduced hip range of motion (ROM) over the gait cycle and reduced maximal hip extension at terminal stance. *Knee flexion/extension* was elevated in 17 participants, bilaterally (PT 2, 5, 6, 7, 10, 13, 15, 16, 18) or unilaterally (PT 1, 3, 4, 9, 12, 14, 17). A recurrent feature was reduced knee flexion during the swing phase, present in 12 participants. Increased flexion at midstance was observed in 3 participants (PT 2, 13, 15), while 4 others (PT 6, 12, 14, 18) leaned towards knee hyperextension at midstance. One outlier (20.9°) (PT 10) was identified for knee flexion/extension at the left side only, corresponding to an overall reduced knee ROM throughout the gait cycle. Eleven participants showed elevated *ankle plantar/dorsiflexion* scores, bilaterally (PT 5, 9) or unilaterally (PT 3, 10, 11, 12, 13, 14, 16, 17, 18), and 4 of them (PT 3, 9, 10, 11) had a reduced second ankle rocker in common. Both on the left and right side, 1 extreme outlier was identified (PT 5), corresponding to a toe-walking pattern with increased plantar flexion throughout the gait cycle. GPS-scores per participant and reference-data can be found in eTable 3 (in the supplemental files). A boxplot of the total GPS, GPS per plane of motion and MAP is presented in figure 1 A and B. eFigure 1 and 2 in the supplemental files represent the kinematic curves for hip and knee flexion/extension, ankle plantar/dorsiflexion and FPA.

**Figure 1:**
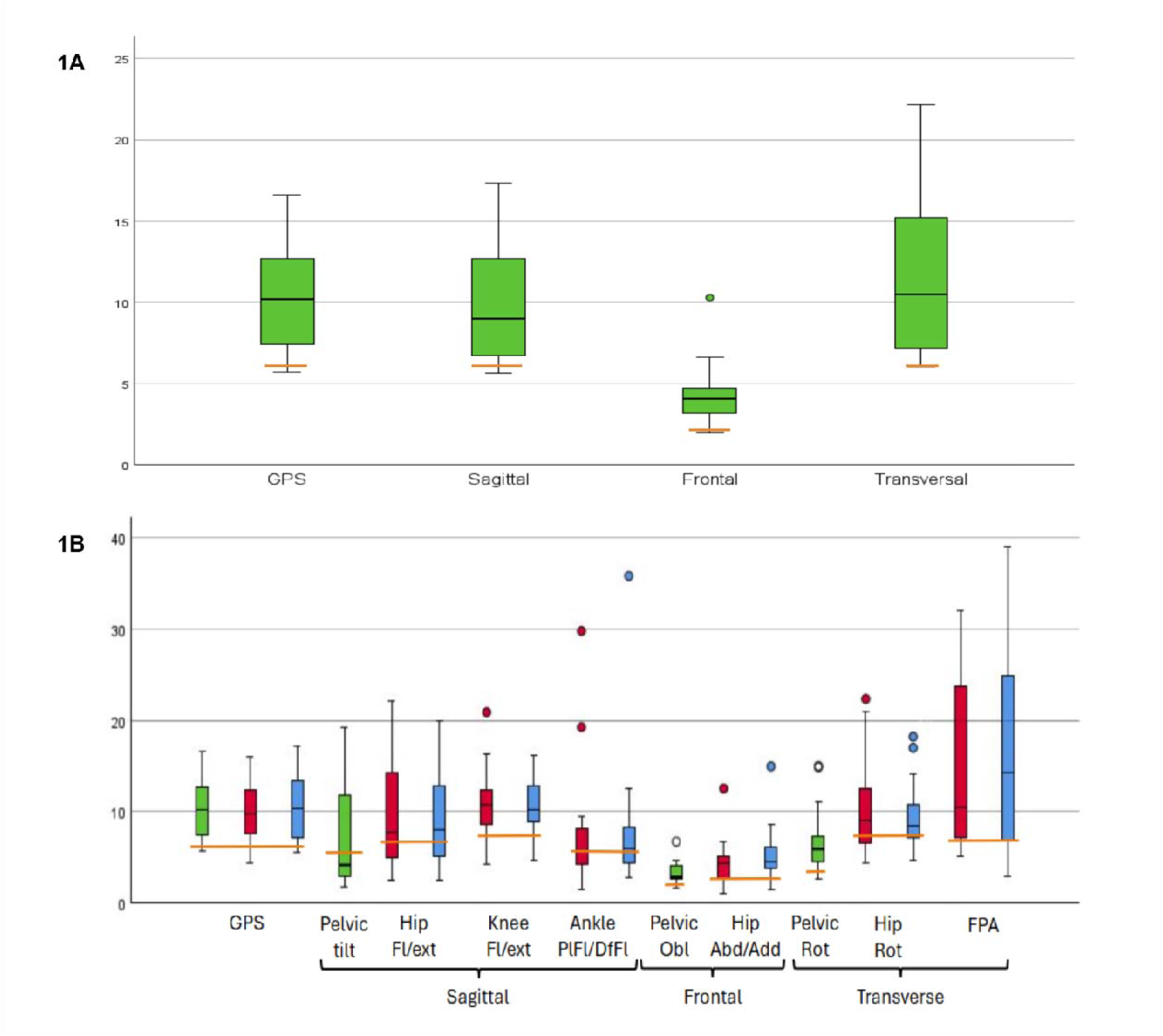
A. Boxplot of the total Gait Profile Score (GPS) and per plane of motion (frontal, sagittal, transversal). B. Boxplot of the Gait Profile Score (GPS) and Movement Analysis Profile (MAP). The green boxplots are the scores of both limbs together. The red boxplots represent the left limb, the blue boxplot the right limb. The dots are the outliers. The orange bars represent the cut-off for normal gait for GPS and MAP, i.e. P95 of the reference dataset.

**Figure 2:**
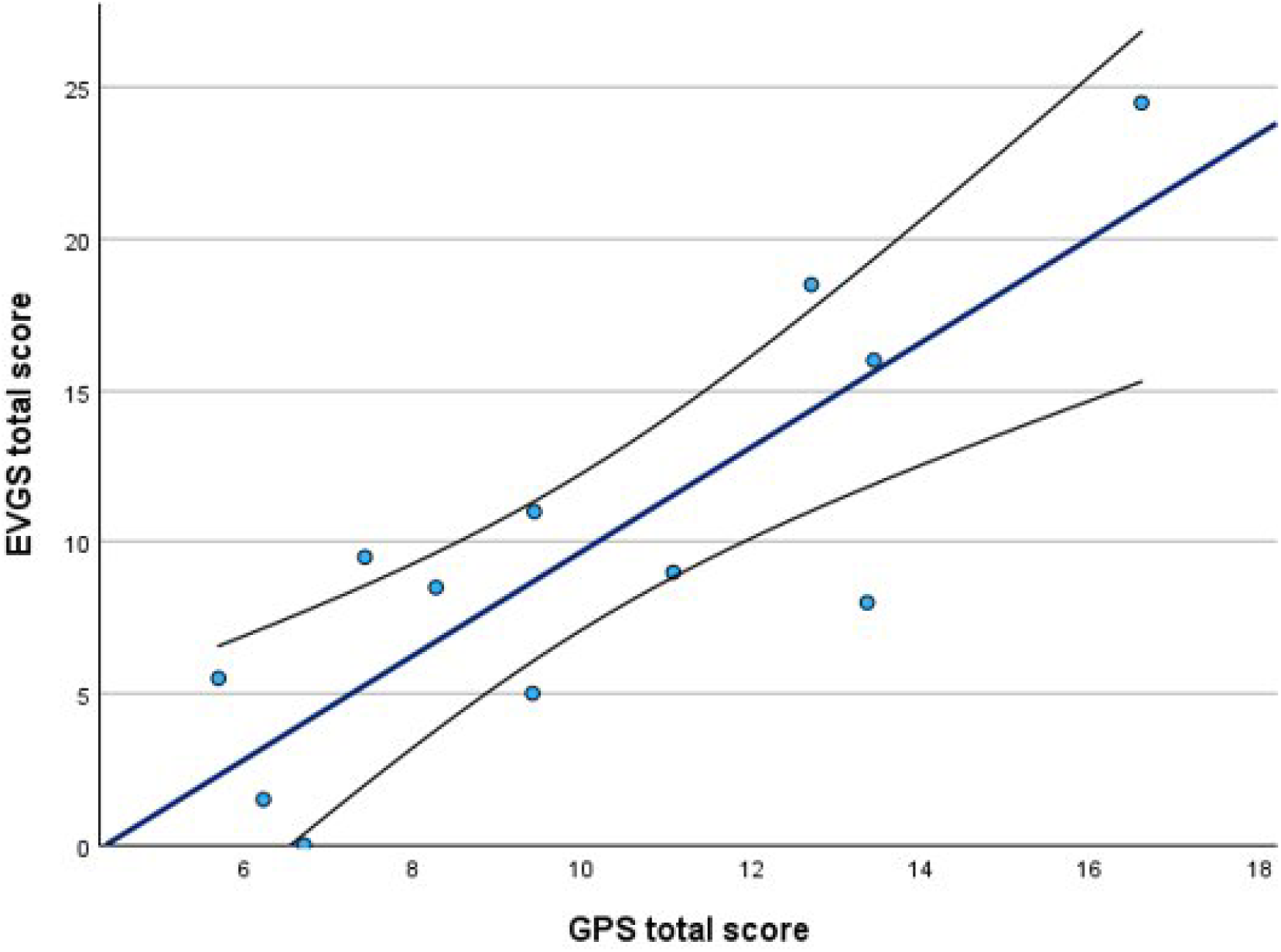
A scatterplot with a fitted linear regression line and a 95% confidence interval of the GPS and EVGS (n = 12).

### 3.3 Functional mobility

Figure 3 displays the results of the FMS-5, FMS-50 and FMS-500, available for 16/18 participants. In the home setting (FMS-5), only 1 participant (PT10) did not walk independently. Remaining participants walked independently without aids on all (n=6) or level surfaces (n= 9). In the community (FMS-500), mobility was more variable: 2/16 participants walked without aids on any surface, 9/16 were independent on flat surfaces, 1/16 needed one or two sticks, 1/16 used a walker and 3/16 used a wheelchair. These findings demonstrated increasing mobility limitations with longer distances and in more complex environments.

**Figure 3:**
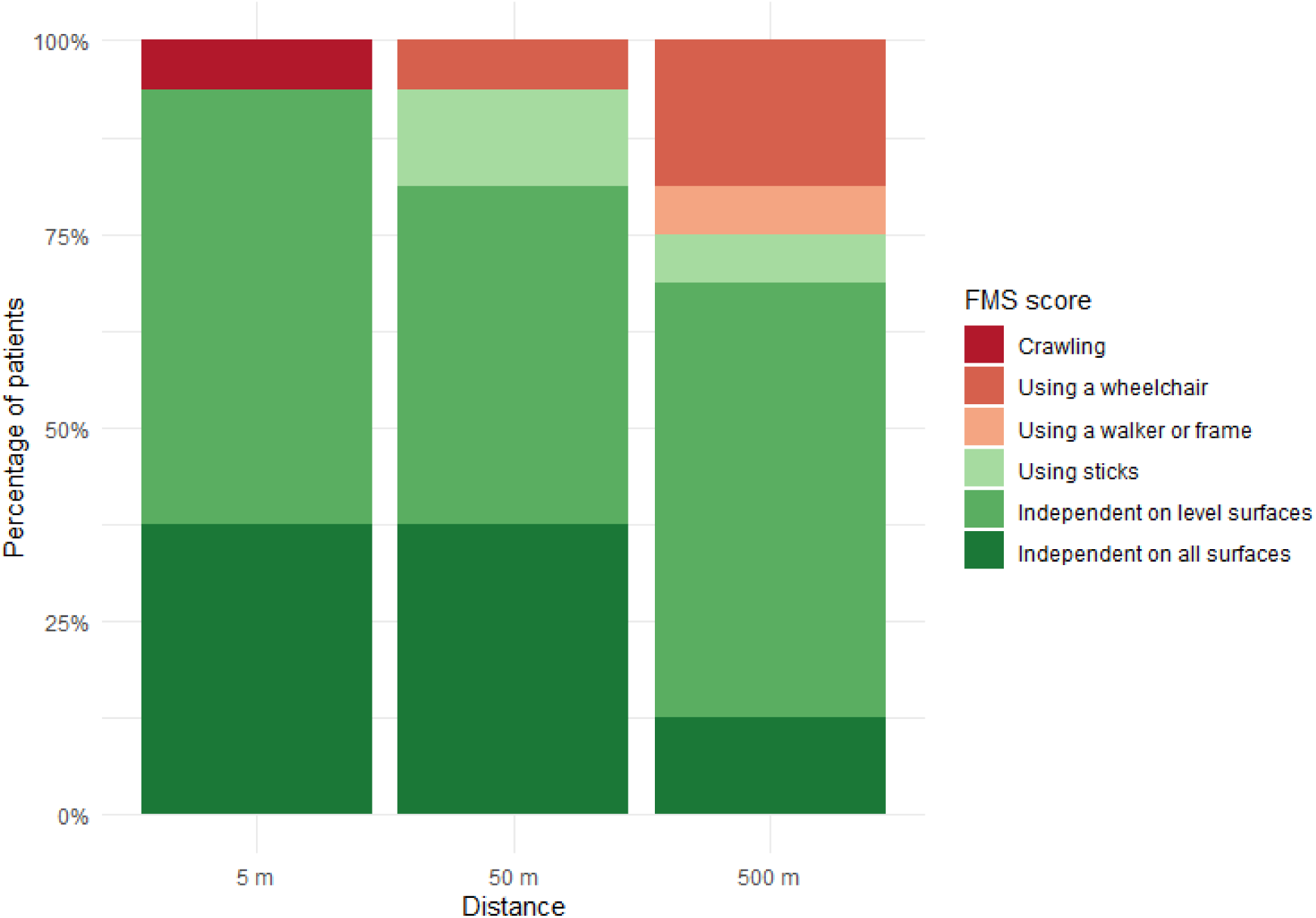
Functional Mobility Scale (FMS) per walking distance: in home setting (5m), in school setting (50m), in community setting (500m), expressed as the percentage of participants (n = 16).

MobQues28-total scores, available for 17/18 participants, ranged from 14.29 to 95.54 with a median score of 57.14 (IQR 32.59 - 82.14). Figure 4 summarizes MobQues28 ratings per item. Overall, caregivers reported no or only slight difficulties in basic activities such as walking indoors/outdoors barefoot or shod on different surfaces, consistent with the FMS, standing still barefoot or shod, getting up from and sitting down on a chair. In contrast, over 50% of the cohort reported significant difficulties in performing following activities: bending down to the floor, getting out of the shower or car, walking down the stairs (with something in the hands), walking over obstacles and running on different surfaces.

**Figure 4:**
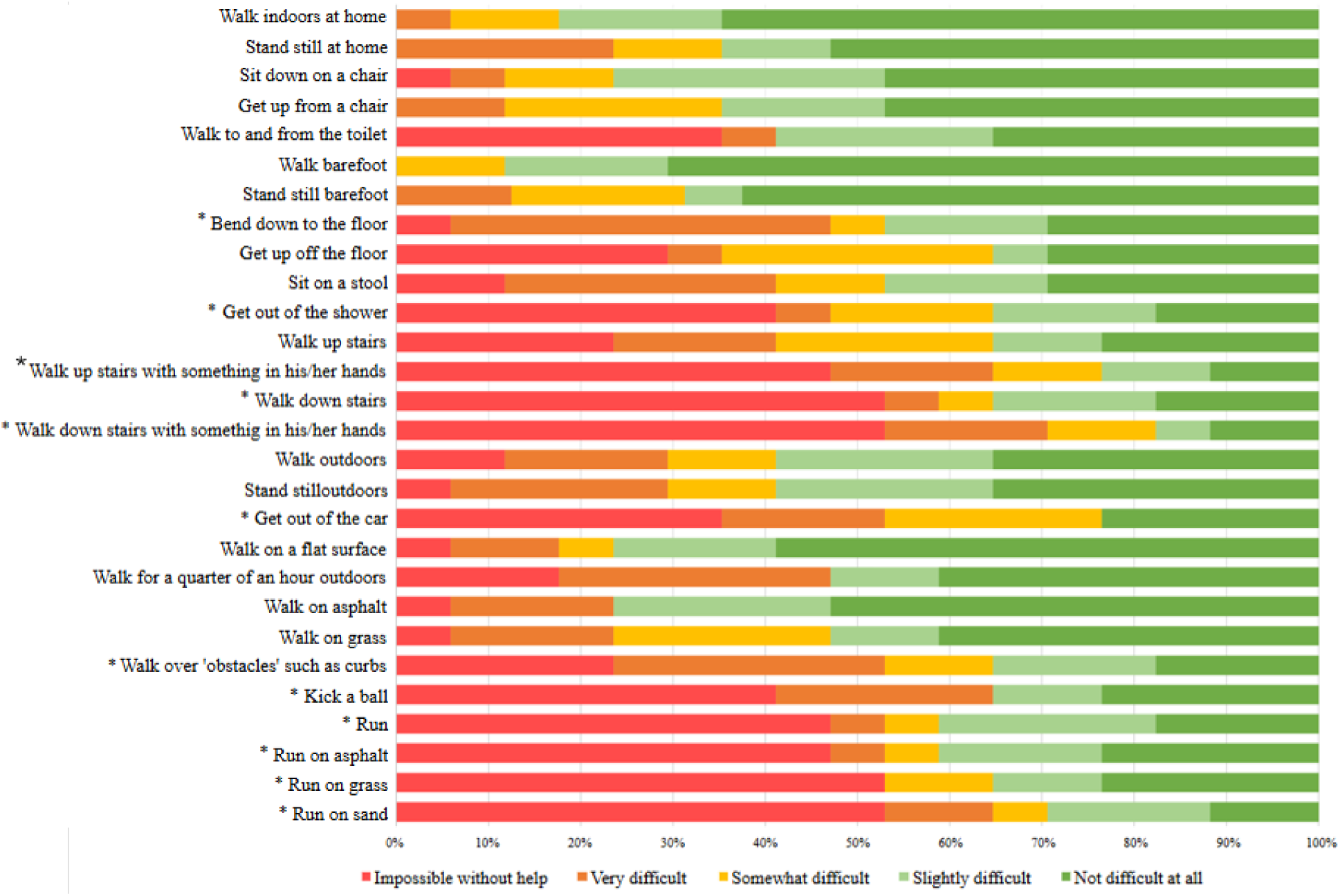
Caregiver responses on the Mobility Questionnaire 28 (MobQues28) within the study cohort (n= 17) in a 100% stacked bar format. Each bar represents the relative contribution of answer categories (Impossible without help; Very difficult; Somewhat difficult; Slightly difficult; Not difficult at all) for one item. *50% or more of study cohort scored ‘Impossible without help’ or ‘Very difficult’.

### 3.4 Caregiver quality of life

PedsQL-FIM-total scores, available for 17/18 participants, ranged from 23.61 to 77.78 with a median score of 60.42 (IQR 53.82 - 66.32). Figure 5 displays an overview of the PedsQL-FIM ratings per item. More than 50% of caregivers reported no or only very little difficulties in the categories family relationships (e.g., conflicts between family members) and cognitive functioning in their caregiving role. However, 50% or more reported that family activities consistently required more time and effort, and reported frequent worry about their child’s future.

**Figure 5:**
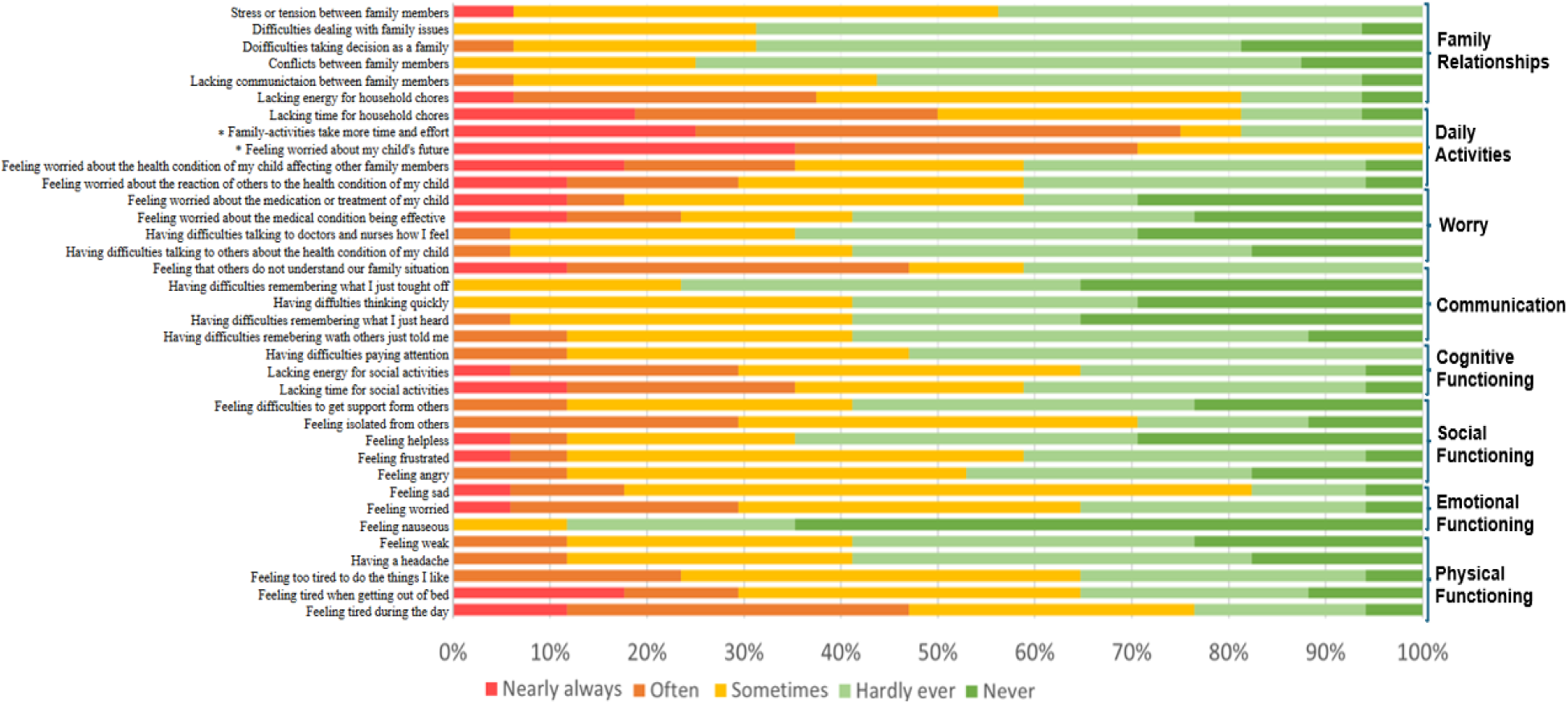
Relative contribution for every item-score on the PedsQL-Family Impact Module (PedsQL-FIM) within the study cohort (n=17). Each bar represents the relative contribution of answer categories (Nearly always; Often; Sometimes; Hardly ever; Never) for one item. *50% or more of study cohort scored ‘Nearly always’ or ‘Often’.

### 3.5 The relation between GPS, functional mobility, genetic and clinical characteristics

A moderate negative and high negative, but not statistically significant, correlation was found between GPS and FMS-500 and between GPS and MobQues28, respectively.

When further evaluating the relation between gait deviations and genetic and clinical characteristics, no significant correlations were identified between GPS and the (1) STXBP1 variant type, (2) age at independent walking, (3) degree of ID, (4) age at seizure onset and (5) seizure frequency.

When looking at the relation between functional mobility, as assessed with the FMS-500, and the same set of characteristics, a high negative, statistically significant correlation was identified between FMS-500 and age of independent walking, meaning that younger age at onset of independent walking correlated with better community mobility. None of the other characteristics showed a significant correlation with FMS-500 after Bonferroni correction. A summary of the statistical analyses comparing GPS and FMS-500 with genetic and clinical characteristics can be found in table 2.

**Table 2:** Results of statistical analyses. In bold are the significant results after Bonferroni correction (α’ = 0.0125 for Mann*-*Whitney U tests and α’ = 0.0083 for Spearman rank correlations).

| Variables | n | Test statistics<br>(*r* /*U*/ *ρ*) | *p*-value |
| --- | --- | --- | --- |
| GPS and EVGS | 12 | *r* = 0.839 | 0.001<sup>1</sup> |
| GPS and functional mobility |  |  |  |
| GPS and FMS-500 | 16 | *ρ* = -0.467 | 0.068<sup>2</sup> |
| GPS and MobQues28 | 17 | *r* = -0.425 | 0.089<sup>1</sup> |
| GPS and clinical characteristics |  |  |  |
| GPS and genetic variant | 18 | *U* = 27.0, *Z* = -1.024 | 0.328<sup>3</sup> |
| GPS and ID | 18 | *U* = 11.0, *Z* = -1.805 | 0.079<sup>3</sup> |
| GPS and frequency of seizures | 18 | *ρ* = -0.154 | 0.542<sup>2</sup> |
| GPS and age at seizure onset | 13 | *ρ* = -0.443 | 0.130<sup>2</sup> |
| GPS and age of independent walking | 17 | *ρ* = 0.389 | 0.122<sup>2</sup> |
| FMS-500 and clinical characteristics |  |  |  |
| FMS-500 and genetic variant | 16 | *U* = 16.0, *Z* = -1.816 | 0.114<sup>3</sup> |
| FMS-500 and ID | 16 | *U* = 13.0, *Z* = -1.477 | 0.21<sup>3</sup> |
| FMS-500 and frequency of seizures | 16 | *ρ* = 0.308 | 0.245<sup>2</sup> |
| FMS-500 and age at seizure onset | 13 | *ρ* = 0.676 | 0.016<sup>2</sup> |
| FMS-500 and age of independent walking | 16 | *ρ* = -0.842 | **<0.001<sup>2</sup>** |
| <sup>1</sup>Pearson correlation, <sup>2</sup>Spearman Rank correlation, <sup>3</sup>Mann-Whitney *U* test |  |  |  |

### 3.6 Correlation i3DGA and EVGS

For the Antwerp cohort (n=12), video-images taken during gait analyses were scored using the EVGS. EVGS showed a high positive correlation with GPS, indicating that higher GPS scores, representing greater deviations from typical gait kinematics, correlated with higher EVGS scores, reflecting more pronounced observable gait abnormalities (Figure 2, Table 2). No ceiling or floor effects were observed; only 1 participant had the minimum score (8.33% of participants, threshold: 15%) and no one reached the maximum score. The scores also did not cluster near the minimum or maximum value.

## 4. DISCUSSION

Prior systematic and quantitative research on gait deviations in STXBP1-RD is scarce. In this cohort, gait of individuals with STXBP1-RD was characterized by reduced walking speed, with shorter step and stride length than typically developing peers. No single pathognomonic kinematic gait pattern was identified with i3DGA. Instead, gait was heterogenous, with an externally rotated FPA as the most recurrent deviation in 11/18 participants. In the sagittal plane, knee kinematics showed marked variability, including both excessive knee flexion as hyperextension during midstance, along with reduced flexion during swing phase. A decrease in overall hip range of motion and especially in peak hip extension at terminal stance were observed. The reduced hip extension in stance and diminished knee flexion in swing, may partially relate to slower walking speed, as described in previous studies (27). These variable finding contrast with Dravet syndrome, where more characteristic crouch-like patterns have been described, and they underscore the heterogeneity of gait abnormalities in STXBP1-RD (12, 13). The frontal MAP showed least variability which might be attributed to this MAP subscore only containing 2 variables, compared to 4 in the sagittal and 3 in the transverse plane of motion. In addition, pelvic obliquity and hip ab-adduction typically show a small range of motion in normal gait. On group level, variation in GPS in our STXBP1 cohort was not significantly different from the normal population, which is likely explained by the large variability and the small cohort size.

Notably, the externally rotated FPA and the aberrant midstance knee patterns merit clinical attention. From a biomedical perspective, an external FPA alters the distribution of foot pressure and the balance in loading of the medial and lateral side of the leg, potentially causing or worsening pes planovalgus. External foot progression is often related to rotational deformities, which, when left untreated, can lead to lever-arm disfunctions affecting hip and knee function as well as progressive foot deformity (28, 29).

Increased or sustained knee flexion during various phases of the gait cycle, in turn, increases joint loading (30), impairs muscle function (31) and increases energy demands (32). These biomechanical alterations create a cycle of worsening function (31) negatively affecting functional mobility and independence.

Knee hyperextension is a heterogeneous, population-specific phenomenon. In children with knock-knee, it coincides with shorter stride length and slower walking speed, patterns that are also evident in our STXBP1-RD cohort (33). Although knee flexor weakness and gastrocnemius spasticity may contribute, dysfunction of the ankle plantar flexors appears to be the primary driver of stance-phase hyperextension (34, 35). These risk factors merit assessment in individuals with STXBP1-RD.

Looking at functional mobility in this cohort, the FMS showed that although almost all patients (93.75%) in this cohort walked independently at least on level surfaces at home, only 37.50% walked independently on all surfaces. Community-level ambulation was even more constrained, with some requiring aids or a wheelchair. FMS findings were mirrored by the results of the MobQues28, indicating more difficulty with increasing task complexity and environmental challenges.

Importantly, our study included ambulatory participants only, which explains the significantly better outcomes in FMS compared to our previous work (5). As such, these results do not reflect the full spectrum with regards to functional mobility in STXBP1-RD.

Previous research showed that patients with no history of seizures had better GMFCS scores and were 4 times more likely to be able to walk (7). In addition, multiple studies have shown that patients with later age of seizure onset are more likely to have better motor outcomes, including a higher likelihood of walking independently (5–7). In this study cohort, we did not find a relationship between age at seizure onset nor seizure frequency and the severity of gait problems or limitations in functional mobility. We hypothesize this is largely due to the small cohort and exclusion of non-ambulatory individuals in our cohort.

Our analysis did show a negative correlation between the age at onset of independent walking and the functional mobility outcome. This is consistent with findings previously reported in a gait analysis study of patients with DS (14). Consequently, the age at which patients acquire the skill to walk independently may serve as a meaningful predictor of functional mobility later in life.

Finally, to explore the impact of impairments in functional mobility on caregiver QoL we used the PedsQL-FIM. The results in this study cohort showed no or only very little difficulties in the categories family relationships and cognitive functioning. However, even in this restricted subset of ambulatory patients, 50% or more of the caregivers reported that family activities consistently required more time and effort. Furthermore, > 50% reported to often or always worry about their child’s future. Previous research on STXBP1-RD and cerebral palsy has shown that mobility may contribute, alongside other biopsychosocial factors, to the overall disease burden of families (10, 11). Understanding the needs and burden associated with this disease is essential for developing more tailored, person-centred support for families, including targeted approaches to motor rehabilitation.

Although GPS is considered the gold standard for quantifying gait deviations, as mentioned before, it is not practical for routine clinical use. In this study, a high positive correlation was observed between GPS and EVGS, which corresponds to previous research (19, 38). EVGS is easier to administer and may serve as a semi-quantitative alternative to i3DGA. It is more discriminative then GMFCS, which categorizes ambulant patients only as level I or II, thereby EVGS has a higher ability to detect subtle differences. Even within this small cohort, EVGS demonstrated a wide range of scores, without ceiling or floor effect. Nevertheless, further evaluation of its sensitivity and discriminative capacity is required before implementation.

## 5. LIMITATIONS AND FUTURE RESEARCH

This study has limitations. First, the cohort size is modest, which reflects both the rarity of the disorder and need for specialized i3DGA facilities at tertiary centers. Consequently, generalizability and statistical power for detecting more subtle associations with clinical features are limited. Second, only ambulatory individuals were included, which does not capture the full spectrum of functional mobility in STXBP1-RD and may attenuate associations with seizure variables and GPS at the group level. Third, although the two sites harmonized marker sets and processing, EVGS was only scored in a subset of participants due to data-sharing restrictions, so generalization of the EVGS-i3DGA correlation requires further validation. Finally, the cross-sectional design precludes inferences about longitudinal change and treatment response.

Despite these limitations, this study provides a quantitative foundation for gait characterization in STXBP1-RD, identifies externally rotated FPA as a common feature, and highlights early independent walking as a clinically meaningful predictor of later community mobility. Collectively, these findings inform clinical decision-making by highlighting the need for careful monitoring of foot alignment in individuals with external foot rotation gait patterns and appropriate referral to physiotherapy or orthopaedics when indicated, while also guiding the selection of scalable outcome measures for natural history studies and interventional trials. Given the strong EVGS-i3DGA correlation, EVGS may be a practical assessment option for broader multicenter cohorts, provided that inter-rater reliability is established.

Future research should extend these results in larger, more diverse cohorts with longitudinal follow-up, as planned in the ongoing European natural history study (NCT06625112). Developing and validating scalable, seizure-independent outcome measures, including motor assessments, will be essential for supporting clinical trial readiness and developing disease-modifying therapies. In addition, incorporating physiotherapeutic assessments into routine clinical care may improve the characterization of externally rotated FPA and sagittal knee deviations, facilitate identification of underlying structural deformities or muscle weakness, and guide individualised interventions including orthopaedic insoles or targeted physiotherapy.

## Data Availability

All data produced in the present study are available upon reasonable request to the authors

## 6. ACKNOWLEDGMENTS AND FUNDING

The authors thank the patients and their families for contributing to this study. They further acknowledge their collaborators of the European STXBP1 consortium for patient referral. This work was supported by a research grant from the University of Pennsylvania Orphan Disease Center in partnership with the Lulu’s Crew/STXBP1 Disorders. SW is a member of the µNEURO Research Centre of Excellence of the University of Antwerp and received funding from FWO (1861424N) and the Queen Elisabeth Medical Foundation (UCB Award).

